# The impact of host resistance on cumulative mortality and the threshold of herd immunity for SARS-CoV-2

**DOI:** 10.1101/2020.07.15.20154294

**Authors:** José Lourenço, Francesco Pinotti, Craig Thompson, Sunetra Gupta

## Abstract

The risk of severe disease and death from COVID-19 is not uniformly distributed across all age classes, with the bulk of deaths occurring among older ages and those with comorbidities. Evidence is also mounting that some individuals have pre-existing immune responses to SARS-CoV-2 which may confer resistance to infection. We present a general mathematical framework which can be used to systematically explore the impact of variation in resistance to severe disease and infection by SARS-CoV-2 on its epidemiology. We find that the herd immunity threshold (HIT) can be lowered by the existence of a fraction of the population who are unable to transmit the virus, whether they are effectively segregated from the general population or mix randomly. These results help to explain the wide variation observed globally in seroprevalence and cumulative deaths and raise the possibility that the proportion exposed may have already exceeded HIT in certain regions.

## Introduction

The spread of a novel infectious agent eliciting protective immunity is typically characterised by three distinct phases: (I) an initial phase of slow accumulation of new infections (often undetectable), (II) a second phase of rapid growth in cases of infection, disease and death, and (III) an eventual slow down of transmission due to the depletion of susceptible individuals, typically leading to the termination of the first epidemic wave. The point of transition between phases I and II is known as the herd immunity threshold (HIT), which is defined as the level of exposure at which an epidemic growth rate becomes negative and incidence starts to decrease. Subsequent epidemics will only occur when the susceptible population is replenished sufficiently through births and/or loss of immunity. Such susceptible-infected-recovered (SIR) systems eventually settle to an endemic equilibrium where the proportion immune is maintained at the HIT with no net change in numbers infected. In practice, most SIR systems tend to exhibit small oscillations around this equilibrium (e.g. due to seasonal effects), leading to regular annual or extra-annual peaks, possibly accompanied by an increase in deaths among those who are naive to infection or experiencing immune senescence. In this endemic state, those who are naive to infection typically comprise young children who have been born subsequent to the last epidemic.

Prior to the first epidemic wave, however, the entire population is expected to be naive to infection. Yet the bulk of deaths from COVID-19 has occurred among older age classes and those with comorbidities ^1,2^, indicating that some level of pre-existing resistance to severe disease may be lost through immune senescence or in association with certain clinical conditions. T-cell and IgG antibody activity have been reported in non-exposed individuals to SARS-CoV-2, suggesting that resistance to disease, and even infection, may accrue from previous exposure to endemic corona viruses ^3–6^. A fraction of the population may also already be intrinsically resistant to infection as a consequence of high functioning innate immunity and such mechanistic reasons as reduced expression of Angiotensin Converting Enzyme 2 (ACE2) ^3,7–11^. Here we present a general framework to assess whether these observations have a significant impact on the epidemiology of SARS-CoV-2 and on our interpretation of mortality, infection and seroprevalence data pertaining to the ongoing pandemic.

## Results

Our model (see **Supplementary Text File 1**) links two subpopulations (groups 1 and 2) by means of an interaction matrix in which δ (0 < δ < 1) specifies the degree of within-group mixing in a subpopulation of proportion ρ (group 1). Thus, all contacts are within the respective groups (i.e. mixing is fully assortative) when δ = 1, and between-group mixing is maximised at δ = 0. Random or proportionate mixing occurs when δ = ρ. We define the basic reproduction number (*R*_0_) for each group as the fundamental transmission potential of the virus within a homogenous population consisting of members of that group. Rates of loss of infection and immunity are given respectively as σ and γ.

If a fraction ρ of the population is resistant to infection 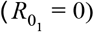, the herd immunity threshold (HIT) is given as 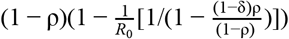, for all possible rates of loss of immunity γ. This suggests that a wide variation in HIT can be observed depending on the proportion resistant ρ, the *R*_0_ within the non-resistant group and the degree of mixing between the resistant (group 1) and non-resistant (group 2). When mixing is fully assortative (δ = 1), HIT = (1 − ρ)(1 − 1/*R*_0_). In other words, the HIT declines in proportion to the size of the resistant group (**Figure 1A**). For example, when *R*_0_ = 2, HIT will be reached at 25% if half the population is resistant. By contrast, under proportionate mixing (δ = ρ, i.e. random mixing), HIT = 1 − 1/*R*_0_ – ρ (**Figure 1B**). This implies that the pathogen will not spread unless the proportion immune is below 1 − 1/*R*_0_. Thus, under the same condition of half the population being resistant, no major epidemic is expected unless *R*_0_ > 2. The HIT determines the level of exposure at which an epidemic growth rate becomes negative and incidence starts to decrease. However, dependent on the level of infection when growth becomes negative, final exposure (**Figures 1 C & D**) is always expected to overshoot the HIT (**Figures 1 E & F**).

**Figure 1.**
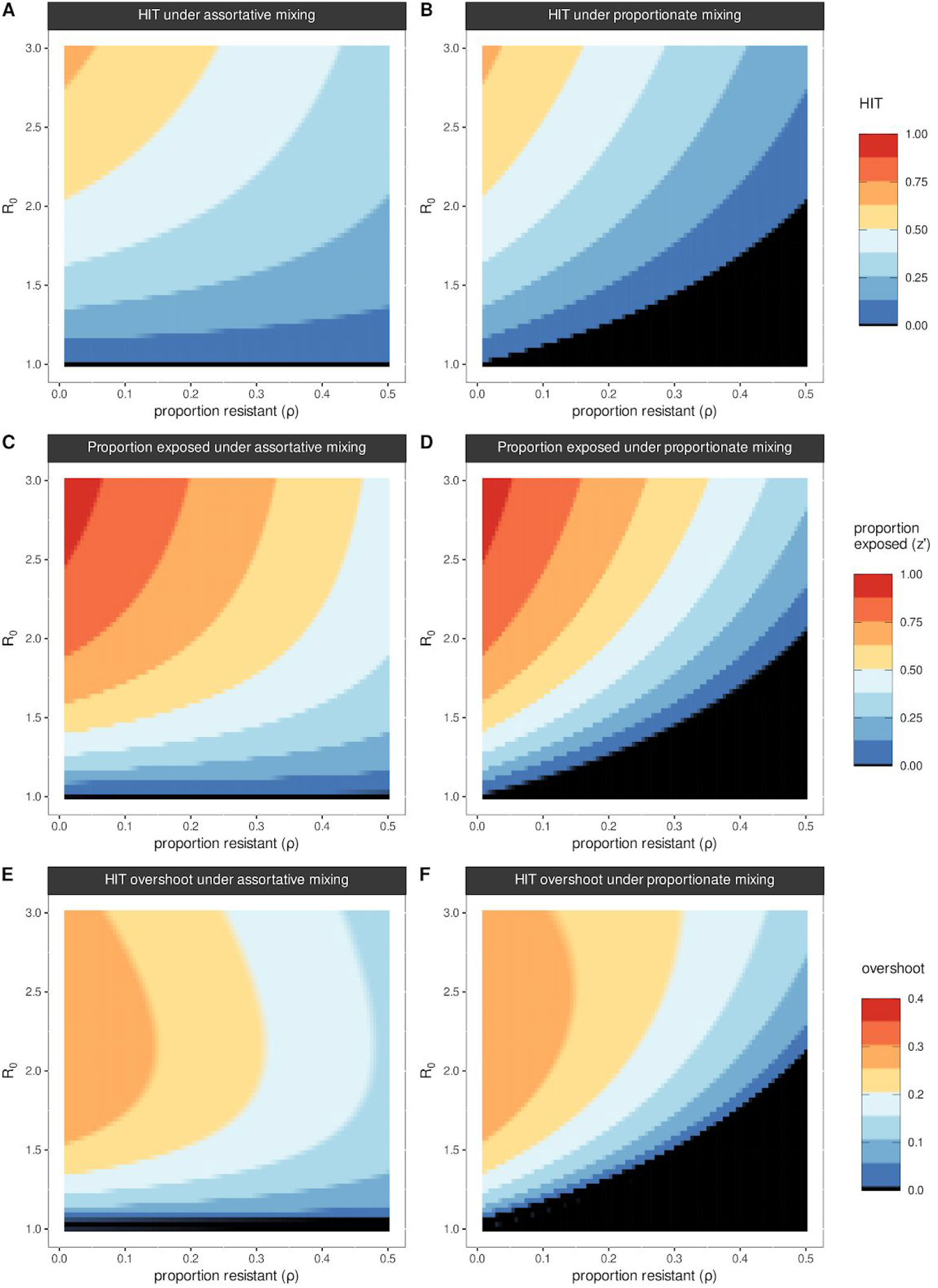
Herd immunity threshold under proportionate and assortative mixing. Herd immunity threshold (HIT) when varying proportion of the population fully resistant (ρ) under (A) assortative mixing (δ = 1), (B) proportionate mixing (δ = ρ). (C) Proportion exposed (z’, i.e. final exposed at the end of simulation) under proportionate mixing, (D) same as panel C but for assortative mixing. (A-D) Each color band equates to a ∼0.125 change, and black covers the range 0-0.01. (E) HIT overshoot (i.e. final exposed at the end of simulation z’ minus HIT), under proportionate mixing, (F) same as in panel E but for assortative mixing. (E-F) Each color band equates to a ∼0.05 change, and black covers the range ∼0-0.01. (A-F) 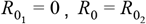, 1/σ = 5 days, γ = 0.

The dependence of HIT on the degree of within-group mixing (δ) increases with the proportion resistant ρ (**Figures 2 A & C**), exhibiting their lowest values in a disassortative extreme (δ = 0). Substantial reductions in cumulative mortality can thus be obtained as the resistant proportion increases (**Figures 2 B & D**) which could provide a simple explanation for the wide variation in death rates reported across various regions. Incomplete resistance can be implemented within this framework by allowing 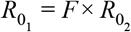 where 0 < *F* < 1 (**Figure S1**). Our simulations indicate that under incomplete resistance, the reduction in HIT is roughly proportional to *F* (for example, 50% of the population being 50% resistant is roughly equivalent 25% with complete resistance, under proportionate mixing, when *R*_0_ = 1.5).

**Figure 2.**
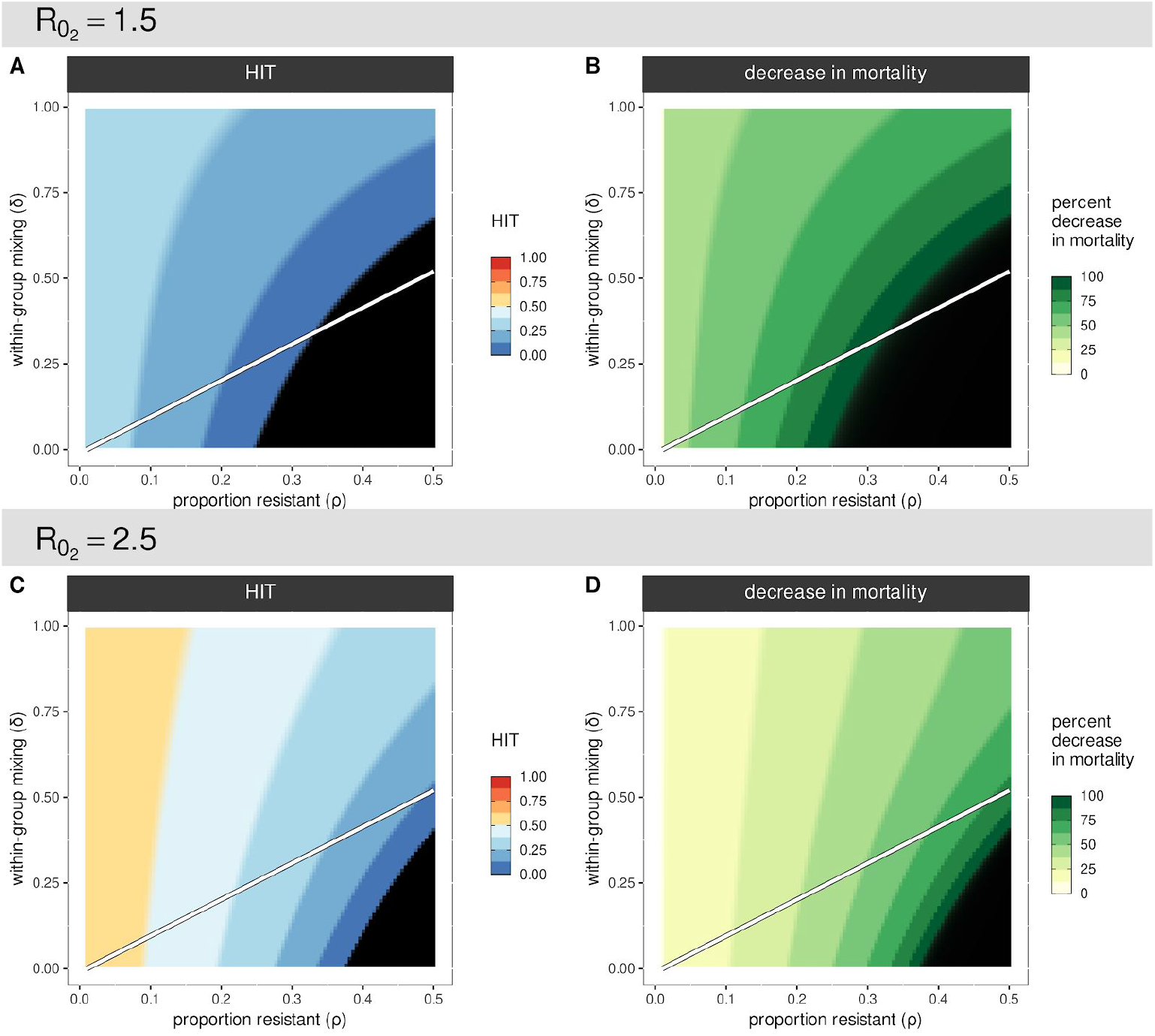
Herd immunity threshold and associated percentage decrease in mortality. Herd immunity threshold (A) and percent decrease in mortality (B) for *R*_0_ = 1.5 under different combinations of proportion resistant (ρ) and levels of within-group mixing (δ). (C-D) The same output as in panels A and B but for *R*_0_ = 2.5. Simulations ran for 365 days with 1/σ = 5 days, γ = 0, 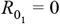 and 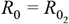 Color bands in the color scales equate to ∼0.125 (A-C) or 12.5 (B-D) change, and black covers the range ∼0-0.01 (A-C) or ∼99-100 (B-D). For visualisation purposes the percent decrease in mortality is 100 × (1 − *z*_*p*_/*z*_ρ=0_), where *z* is the proportion exposed at the end of the simulations with a fraction resistant to infection (*z*_ρ_) and without (*z*_ρ=0_). The white line designates proportionate mixing (ρ = δ) separating an area of assortative (higher within group) mixing above from disassortative (higher between groups) mixing in the area below.

Reductions in cumulative mortality as presented in **Figures 2 B & D** rest on the relative difference (ratio) between the final epidemic sizes of simulated outbreaks with (*z*_ρ_) and without (*z*_ρ=0_) a resistant fraction to infection. However, cumulative mortality can be precisely derived within this general framework by assigning appropriate infection fatality rates to the two subpopulations. For SARS-CoV-2, this can be achieved by factoring in a delay between infection and death, and defining the groups as being resistant and vulnerable to death rather than infection. By fitting cumulative mortality to observed counts and applying estimates of the infection fatality rate to the vulnerable fraction, it is then possible to derive early estimations of epidemiological parameters such as the doubling time, *R*_0_, observation rate and proportion exposed (see **Supplementary Text File 1**).

## Discussion

The risk of infection with any pathogen depends crucially on the proportion of the population currently immune. Non-pharmaceutical interventions preventing the proportion exposed from exceeding HIT will leave the population open to further growth in infections once these measures are eased. However, if the proportion of the population exposed is in excess of the HIT, any subsequent epidemic growth will be mitigated until the susceptible population is replenished through births and / or loss of immunity.

Determining the proportion exposed for SARS-CoV-2 is not possible through tracking clinical cases since the majority of infections are likely to be asymptomatic ^12^, although symptom tracking and other proxies such as excess influenza-like-illness provide a promising alternative route ^13,14^. Obtaining these data through serological surveys has proved to be a challenge, principally due to the variability in both antibody and cellular immune responses among exposed individuals and their kinetics ^15,16^. Reported levels of seroprevalence have not come close to what is believed to be necessary for herd immunity ^16–20^. Our results indicate that a wide variation in reported levels of exposure to SARS-CoV-2 can arise as a result of differences in the proportion of the population resistant to infection (ρ) and mixing between groups. Determining whether HIT has been attained is compromised by the tendency of host-pathogen systems to overshoot HIT **(Figures 1C, D, E, F)**. High levels of seropositivity can arise under a reasonable range of the fraction resistant ρ and *R*_0_ where HIT is nonetheless lower than the proportion of the population already exposed (**Figures 1 E & F**). Equally, seropositivity measures of 10-20% are entirely compatible with local levels of immunity having approached or even exceeded the HIT, in which case the risk and scale of future resurgence is lower than currently perceived.

Several other approaches for estimating SARS-CoV-2 HIT have recently been put forward ^21–26^, all indicating that it is below the classic expectation of 1 − 1/*R*_0_. Those that have taken account of the effect of non-pharmaceutical interventions (e.g. ^22^) typically return values of HIT which are compatible with current seroprevalence estimates and with an endemic state where the risk of death to those vulnerable is on a par with other major respiratory illnesses. The exercises presented in this article do not offer a means to precisely determine the HIT of SARS-CoV-2. Our purpose here is simply to explore the possible impact of pre-existing resistance to infection, for which evidence is steadily mounting ^3–6,8–11^.

It is important to note that HIT is independent of the rate of loss of immunity (γ) although the latter will affect the timing and magnitude of the subsequent epidemic peaks (**Figure S2**). Moreover, the public health impact of subsequent peaks will depend on the degree to which previous exposure reduces severity of disease upon reinfection, and not just whether infection-blocking immunity is lost. Given the considerable body of evidence that exposure to seasonal coronaviruses offers homologous protection against clinical symptoms (e.g. ^11^), it would be reasonable to expect that second and subsequent epidemic waves will result in far fewer deaths than the first wave.

## Data Availability

No data is included.

## Acknowledgements

The authors would like to thank Paul Klenerman, Paul Wikramaratna and Alexander Caccia for useful comments on the manuscript.

## Supplementary Files

Supplementary Figures - List and legends of supplementary Figures S1-3 (support of main text). Supplementary Text File - Model details and complementary results.

## References

1. Richardson, S. et al. Presenting Characteristics, Comorbidities, and Outcomes Among 5700 Patients Hospitalized With COVID-19 in the New York City Area. JAMA (2020) doi: 10.1001/jama.2020.6775.

2. Williamson, E. J. et al. OpenSAFELY: factors associated with COVID-19 death in 17 million patients. Nature (2020) doi: 10.1038/s41586-020-2521-4.

3. Grifoni, A. et al. Targets of T Cell Responses to SARS-CoV-2 Coronavirus in Humans with COVID-19 Disease and Unexposed Individuals. Cell 181, 1489–1501.e15 (2020).

4. Meckiff, B. J. et al. Single-cell transcriptomic analysis of SARS-CoV-2 reactive CD4 T cells. bioRxiv (2020) doi: 10.1101/2020.06.12.148916.

5. Bert, N. L. et al. Different pattern of pre-existing SARS-COV-2 specific T cell immunity in SARS-recovered and uninfected individuals. (2020) doi: 10.1101/2020.05.26.115832.

6. Ng, K. W. et al. Pre-existing and de novo humoral immunity to SARS-CoV-2 in humans. bioRxiv 2020.05.14.095414 (2020) doi:10.1101/2020.05.14.095414.

7. Wu, Z. & McGoogan, J. M. Characteristics of and Important Lessons From the Coronavirus Disease 2019 (COVID-19) Outbreak in China: Summary of a Report of 72 314 Cases From the Chinese Center for Disease Control and Prevention. JAMA (2020) doi: 10.1001/jama.2020.2648.

8. Jing, Q.-L. et al. Household secondary attack rate of COVID-19 and associated determinants in Guangzhou, China: a retrospective cohort study. The Lancet Infectious Diseases (2020) doi: 10.1016/s1473-3099(20)30471-0.

9. Davies, N. G. et al. Age-dependent effects in the transmission and control of COVID-19 epidemics. Nat. Med. (2020) doi: 10.1038/s41591-020-0962-9.

10. Nelde, A. et al. SARS-CoV-2 T-cell epitopes define heterologous and COVID-19-induced T-cell recognition. (2020) doi: 10.21203/rs.3.rs-35331/v1.

11. Sekine, T. et al. Robust T cell immunity in convalescent individuals with asymptomatic or mild COVID-19. (2020) doi: 10.1101/2020.06.29.174888.

12. Hoxha, A. et al. Asymptomatic SARS-CoV-2 infection in Belgian long-term care facilities. Lancet Infect. Dis. (2020) doi: 10.1016/S1473-3099(20)30560-0.

13. Silverman, J. D., Hupert, N. & Washburne, A. D. Using influenza surveillance networks to estimate state-specific prevalence of SARS-CoV-2 in the United States. Sci. Transl. Med. (2020) doi: 10.1126/scitranslmed.abc1126.

14. Menni, C., Sudre, C. H., Steves, C. J., Ourselin, S. & Spector, T. D. Quantifying additional COVID-19 symptoms will save lives. Lancet 395, e107–e108 (2020).

15. Burgess, S., Ponsford, M. J. & Gill, D. Are we underestimating seroprevalence of SARS-CoV-2? BMJ 370, m3364 (2020).

16. Stadlbauer, D. et al. Seroconversion of a city: Longitudinal monitoring of SARS-CoV-2 seroprevalence in New York City. (2020) doi: 10.1101/2020.06.28.20142190.

17. Pollán, M. et al. Prevalence of SARS-CoV-2 in Spain (ENE-COVID): a nationwide, population-based seroepidemiological study. Lancet (2020) doi: 10.1016/S0140-6736(20)31483-5.

18. Stringhini, S. et al. Seroprevalence of anti-SARS-CoV-2 IgG antibodies in Geneva, Switzerland (SEROCoV-POP): a population-based study. Lancet (2020) doi: 10.1016/S0140-6736(20)31304-0.

19. Thompson, C. et al. Neutralising antibodies to SARS coronavirus 2 in Scottish blood donors - a pilot study of the value of serology to determine population exposure. (2020) doi: 10.1101/2020.04.13.20060467.

20. Percivalle, E. et al. Prevalence of SARS-CoV-2 specific neutralising antibodies in blood donors from the Lodi Red Zone in Lombardy, Italy, as at 06 April 2020. Euro Surveill. 25, (2020).

21. Britton, T., Ball, F. & Trapman, P. A mathematical model reveals the influence of population heterogeneity on herd immunity to SARS-CoV-2. Science (2020) doi: 10.1126/science.abc6810.

22. Gomes, M. G. M. et al. Individual variation in susceptibility or exposure to SARS-CoV-2 lowers the herd immunity threshold. medRxiv (2020) doi: 10.1101/2020.04.27.20081893.

23. Friston, K. J., Parr, T., Zeidman, P., Razi, A. & Flandin, G. Dynamic causal modelling of COVID-19 [version 2; peer. (2020).

24. Lipton, A. & Lopez de Prado, M. Mitigation Strategies for COVID-19: Lessons from the K-SEIR Model. (2020) doi: 10.2139/ssrn.3623544.

25. Friston, K., Costello, A. & Pillay, D. Dark matter, second waves and epidemiological modelling. medRxiv (2020).

26. Levitt, M., Scaiewicz, A. & Zonta, F. Predicting the Trajectory of Any COVID19 Epidemic From the Best Straight Line. medRxiv 2020.06.26.20140814 (2020).

