## Supplementary material for "The impact of host resistance on cumulative mortality and the threshold of herd immunity for SARS-CoV-2": Text File 1

#### Supplementary Text File 1 - Model details and complementary results

##### Model

Our model (equations 1-4) links two subpopulations (groups 1 and 2) by means of an interaction matrix in which $\delta$ ($0 < \delta < 1$) specifies the degree of within-group mixing in a subpopulation of proportion $\rho$ with the smaller total number of total contacts (equal to $\rho c_1$, where $c_1$ is the contact rate of that group). Thus, all contacts are within the respective groups (i.e. mixing is fully assortative) when $\delta = 1$, and between-group mixing is maximised at $\delta = 0$. Random mixing occurs when $\delta = \rho c_1/(\rho c_1 + (1 - \rho c_2))$ .

$$\dot{z}_i = \lambda_i(1 - z_i) - \gamma_i z_i \quad (1)$$

$$\dot{y}_i = \lambda_i(1 - z_i) - \sigma_i y_i \quad (2)$$

$$\lambda_1 = \beta_1 c_1[\delta y_1 + (1 - \delta)y_2] \quad (3)$$

$$\lambda_2 = \beta_2 c_2[\frac{(1-\delta)\rho c_1}{(1-\rho)c_2} y_1 + (1 - \frac{(1-\delta)\rho c_1}{(1-\rho)c_2})y_2] \quad (4)$$

The proportion infected (and infectious) is termed $y$ and $z$ (infected plus recovered) is the proportion already exposed; $\lambda_1$ and $\lambda_2$ are the per capita rates of infection of group 1 (of proportion $\rho$) and group 2 (of proportion $1 - \rho$) and is determined by the product of the intrinsic transmission rate $\beta_i$ and contact rate $c_i$ of each group (with $i = 1, 2$) in addition to the distribution of the contacts between the different groups [26]. We define the basic reproduction number ($R_0$) for each group as the fundamental transmission potential of the virus within a homogenous population consisting of members of that group; hence $R_{0_i} = \beta_i c_i/\sigma_i$ where $\sigma_i$ is the group-specific recovery rate and $\gamma_i$ is the group-specific rate of loss of immunity (with $i = 1, 2$).

If a fraction $\rho$ of the population is resistant to infection ($R_{0_1} = 0$), the 'herd' immunity threshold (HIT) is given as $z^* = (1 - \rho)(1 - \frac{1}{R_0}[1/(1 - \frac{(1-\delta)c_1\rho}{c_{2(1-\rho)}})])$, for all values of $\gamma$.

The model was solved numerically in R.

##### Deriving incidence of deaths

Our overall approach rests on the assumption that only a fraction of the population is at risk of death. This proportion is itself only a fraction of the risk groups already well described in the literature [27–31], including the elderly and those carrying critical comorbidities (e.g. asthma). Cumulative death counts ($\Lambda$) are obtained by considering that mortality occurs with frequency $\eta$ (i.e. a small group of proportion $\eta$ exists in the population, whom will experience death upon infection), effectively defining the infection fatality ratio (IFR) as $\eta$. We consider the delay between the time of infection and of death ($\psi$) as a combination of incubation period and time to death after onset of symptoms.

If $\beta_1 c_1 = \beta_2 c_2$, then:

$$\Lambda_t = \eta N z_{t-\psi}$$

We calibrate this model to cumulative reported SARS-CoV-2 associated deaths from the United Kingdom (UK) starting on 05/03/2020 (first death) and ending 15 days later (19/03/2020) to avoid potential effects of local control strategies implemented (for dates of interventions, see [31]). In **Figure 1** we present a summary of model output after being fit to the number of deaths in the UK while setting key parameters to restricted priors well within reported ranges (**Table 1**). The model was able to approximate cumulative death counts (**Figure 1A**) for a wide range of possible IFRs.

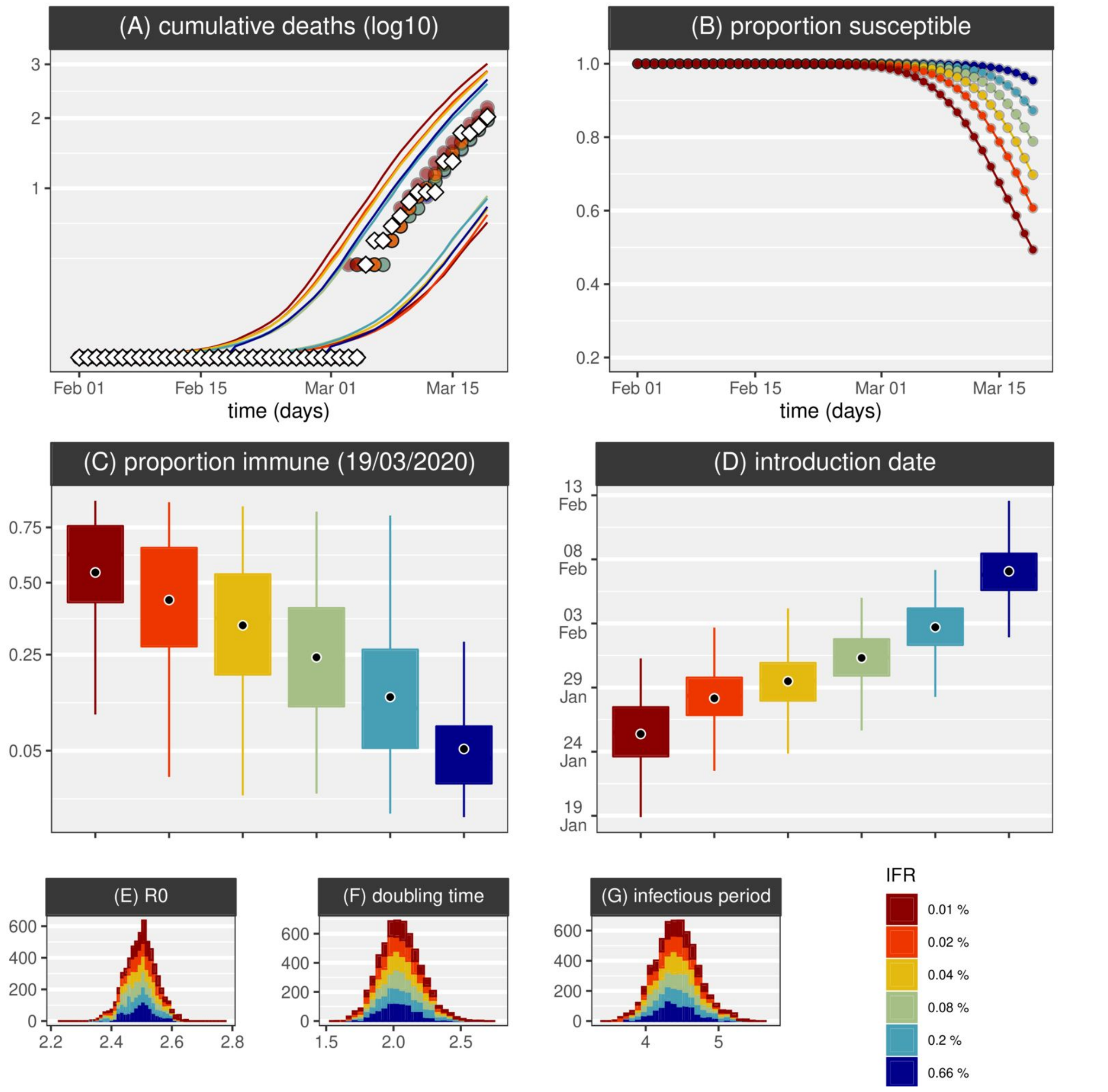


**Figure 1:** Fitting summary for R0=2.5 and various possible IFRs. (A) Data points (black diamonds) and mean model output after fitting (full points) for cumulative mortality. Coloured areas are the model 95% CI. (B) Mean model output after fitting (full points) for the proportion of susceptibles. Model posteriors for (C) time (day) of introduction, (D) herd-immunity on 19/03/2020, (E) R0, (F) doubling time and (G) infectious period. Boxplot hinges present the 25th and 75th percentiles. MCMC ran for 1 million steps. Results presented are the posteriors (model output) using 1000 samples after a burnout of 50%.

As the prior for the IFR is reduced from mean 0.66% to 0.01%, fitting death counts adjusts the introduction date further back to late January, more than a month before the first reported death (**Figures 1D**). As a consequence, accumulated herd-immunity by 19/03/2020 presents an inverse relationship with the assumed IFR prior (**Figures 1C**), with the possibility of thousands of infections having occurred undetected [32–34], even before the first death is reported [35]. Generally, the model respected the priors (**Table 1**) but also adjusted parameters (e.g. **Figure 1G**) required to replicate the necessary doubling times (**Figure 1F**). The latter were estimated to be the same across all considered IFRs (e.g. at 2.07 days, 95% CI 1.78-2.40 for IFR=0.2%) [36,37], well within a range obtained in a sensitivity analysis (**Extra Figure 1**).

A higher $R_0$ (=4) leads to a commensurate increase in the level of herd immunity, but has a minimal effect on the date of introduction; while a longer infection period is required to fit the mortality data but this is still within reported ranges (**Extra Figure 2**). Including an exposed (but not yet infectious) class resulted in a longer doubling time with $R_0 = 2.5$ (**Extra Figure 3**) but accords well with $R_0$ =4 provided infectious periods are lower (**Extra Figure 4**).

Finally we considered the effects of an external contribution to the risk of infection growing exponentially with a doubling time of 5 days from the (estimated) time of first introduction until the date of lockdown (similar to [38], see Methods) to reflect influx of infected individuals into the UK. This had the general effect of delaying our estimates of the date of first introduction by ~7 days (on average) (**Extra Figure 5**). Across such alternative exercises, model output still demonstrated the inverse relationship between the IFR and the proportion currently immune, with the possibility of accumulation of significant levels of herd immunity depending on the IFR considered.

An observation rate ($\theta$, ratio between confirmed cases and total infections) is not explicitly modelled, but can be defined as $\theta = IFR/CFR = (D/I)/(D/C) = C/I$; where D is the number of deaths, I the infections, C the confirmed infections (cases) and CFR the case fatality ratio. From the model's assumed IFR and four CFR independent estimations, we obtained possible distributions for $\theta$ (with introduction of the virus is done as a single event (number of individuals 1/N) at estimated time of introduction $T_i$).

With the true IFR in the UK being unknown, the IFR-dependent scenarios presented (**Figure 1**) remain theoretical projections of both the epidemic duration and accumulation of herd-immunity up to 19/03/2020. We thus looked at contextualizing such projections in light of four previous estimates of the case fatality ratio (CRF) in other epidemiological contexts (see **Methods**). From the relationship $\theta = IFR/CFR$ we obtain the observation rate $\theta$ for each of our modelled scenarios using the respective IFR prior and each of the literature CFRs - effectively considering that any of the reported CFRs could apply to the UK epidemic (**Figure 2**). The CFR considered were: 2.6% (95% CI 0.89-6.7) for the Diamond Princess cruise ship (**Figure 2A**)[39], 3.67% (95% CI 3.56-3.80) for China (**Figure 2B**)[30], 1.2% (95% CI 0.3-2.7) for China (**Figure 2C**)[39], and 1.4% (95% CI 0.9-2.1) for Wuhan (China) (**Figure 2D**)[40]. Although there were significant differences when considering each of the four CRFs (**Figures 2ABCD**), there was a positive relationship between the IFR and the observation rate. The three smallest IFRs modelled ($IFR <= 0.04\%$) were found to be compatible with the reported CRFs when the observation rate $\theta$ was generally close to, or lower than 5% (i.e. 1 or less in 20 infections being confirmed / reported). The $IFR \approx 0.66\%$ is similar to the one recently estimated by Verity and colleagues for China (0.66%, 95% CI 0.39-1.33) [30]. This IFR has been the basis of model projections informing the UK government (see [31,38] for details). In our model (without control) it resulted in mean herd-immunity of 7.05% by 19/03/2020 (**Figure 1C**), estimated introduction on 07/02/2020 (**Figure 1D**), and was compatible with the CFR of the same study for an observation rate of 18.6% (**Figure 2B**).

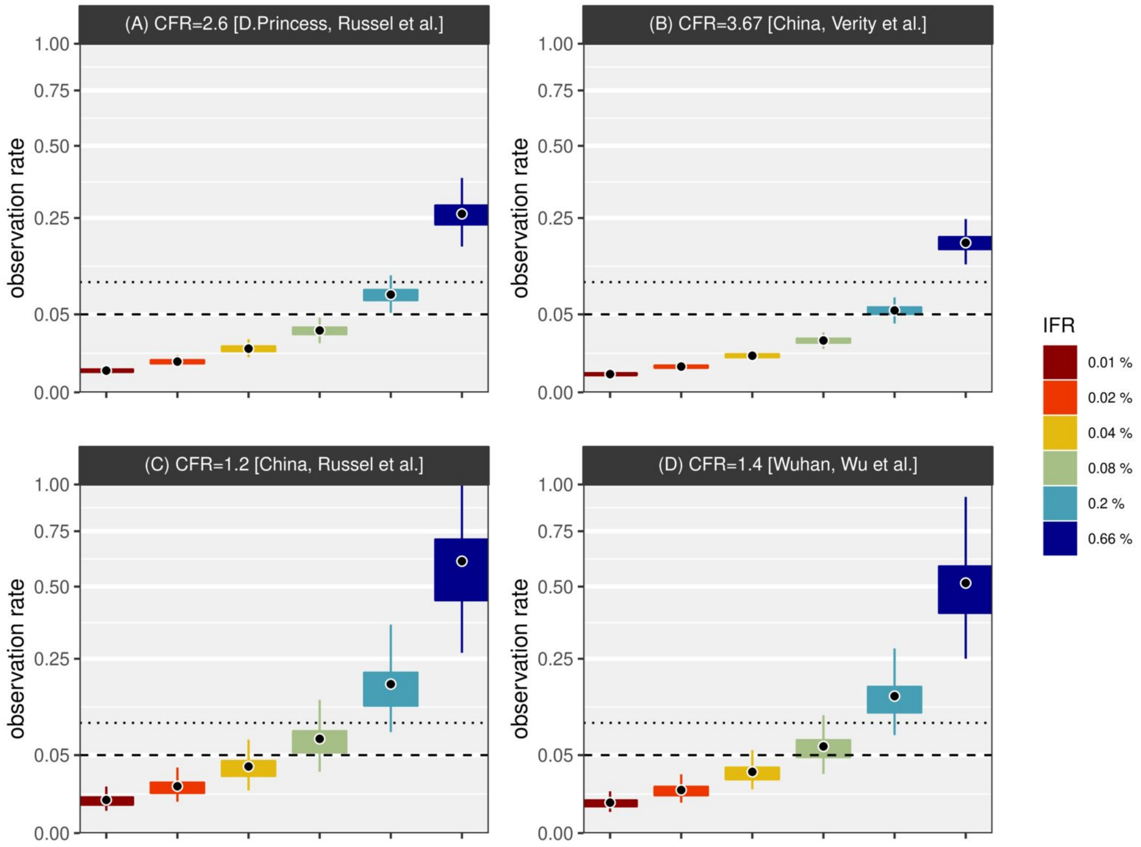


**Figure 2:** Model observation rates for R0=2.5 and various possible IFRs. Four CFR estimations from the literature **(panels A-D**, see main text for references**)** are used to derive model observation rates for each modelled IFR (color legend). Horizontal dotted line marks the 10% observation rate, and dashed line marks the 5% observation rate. Observation rate is the ratio of reported cases and total infections. See Methods for details. Boxplot hinges present the 25th and 75th percentiles. MCMC ran for 1 million steps. Results presented are the posteriors (model output) using 1000 samples after a burnout of 50%.

The UK exercises presented in **Figure 1** for $R_0 = 2.5$ and **Extra Figure 2** for $R_0 = 4$ were repeated for Italy. This resulted in very similar conclusions (**Extra Figures 6-7**), albeit with higher population-level immunity for Italy at 15 days post first reported death.

Overall, these results underscore the dependence of the inferred epidemic curve on the real IFR, showing that accumulation of significant population-level immunity is possible, depending on the currently unknown IFR and observation rate. They also demonstrate how informative the proportion of the population already exposed to SARS-CoV-2 is to determining the IFR.

| **Variable / Parameter** | | **Assumptions / Priors** | **Support** |
|---|---|---|---|
| proportion infectious | $y$ | equation 1 | --- |
| proportion of population no longer susceptible | $z$ | equation 2 | --- |
| cumulative deaths | $\Lambda$ | equation 3 | --- |
| time (day) of introduction | $T_i$ | Uniform distribution (-∞, +∞) | --- |

| | | | |
|---|---|---|---|
| basic reproduction number | $R_0$ | Gaussian distributions<br>G1(M=2.5,SD=0.05)<br>G2(M=4.0,SD=0.05) | 31,41–44 |
| infectious period (days) | $1/\sigma$ | Gaussian distribution<br>1/G1(M=1/4,SD=0.05) | 41,45–47 |
| transmission coefficient | $\beta$ | $\beta = \sigma R_0$ | --- |
| time (days) between symptom onset and death | $\psi$ | Gaussian distribution<br>G1(M=15,SD=1.5) | 45 |
| infection fatality ratio | IFR= $\eta$ | Gaussian distribution<br>$\rho$ = G1(M=m,SD=0.05*m),<br>m $\in \{1/150,\ 1/500, 1/1250\ ...\}$ | --- |
| population size | N | UK 66.87M, Italy 60M | --- |

**Table 1 - Model variables and parameters. M=mean. SD=standard deviation. S=scale. R=rate.**

**Italy:** A time series was obtained from the Italian Department of Civil Protection GitHub repository [48] (accessed on 17/03/2020). We trimmed the data to the first 15 days of death counts above zero (21/02/2020 to 06/03/2020) to include only the initial increase free of effects from local control measures.

**United Kingdom:** A time series was obtained from the John Hopkins University Centre for Systems Science and Engineering COVID-19 GitHub repository [49](accessed 19/03/2020). We trimmed the data to the first 15 days of death counts above zero (05/03/2020 to 19/03/2020) to include only the initial increase free of effects from local control measures.

#### Extra Figures

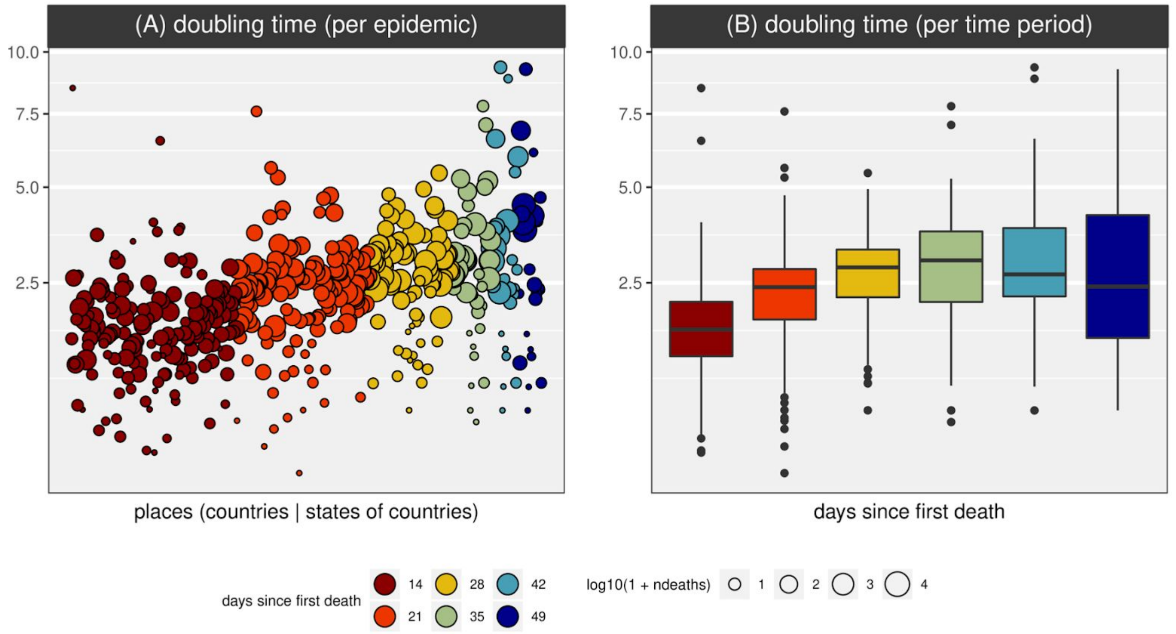


**Extra Figure 1 - Doubling time sensitivity.** Data on cumulative mortality (time series, TS) was downloaded from the Johns Hopkins (JH) Center for Systems Science and Engineering (CSSE) data set [i,ii] on 12 April 2020. For each TS, a growth rate was extracted using *growthcurver* R-package (v0.3), used to calculate a doubling time. Each TS was used to create six new TS starting at the date of first reported death(s) and ending 14, 21, 28, 35, 42 and 49 days after ('days since first death', colour scale). **(A)** Doubling times for all available TS. As more 'days since first death' were considered, a smaller number of TS were available. TS for which obtaining a growth rate was not possible are not shown (e.g. when deaths were constant). **(B)** Summary of panel (A) using boxplots.

[i] https://github.com/CSSEGISandData/COVID-19

[ii] https://systems.jhu.edu/research/public-health/ncov/

[iii] https://CRAN.R-project.org/package=growthcurver

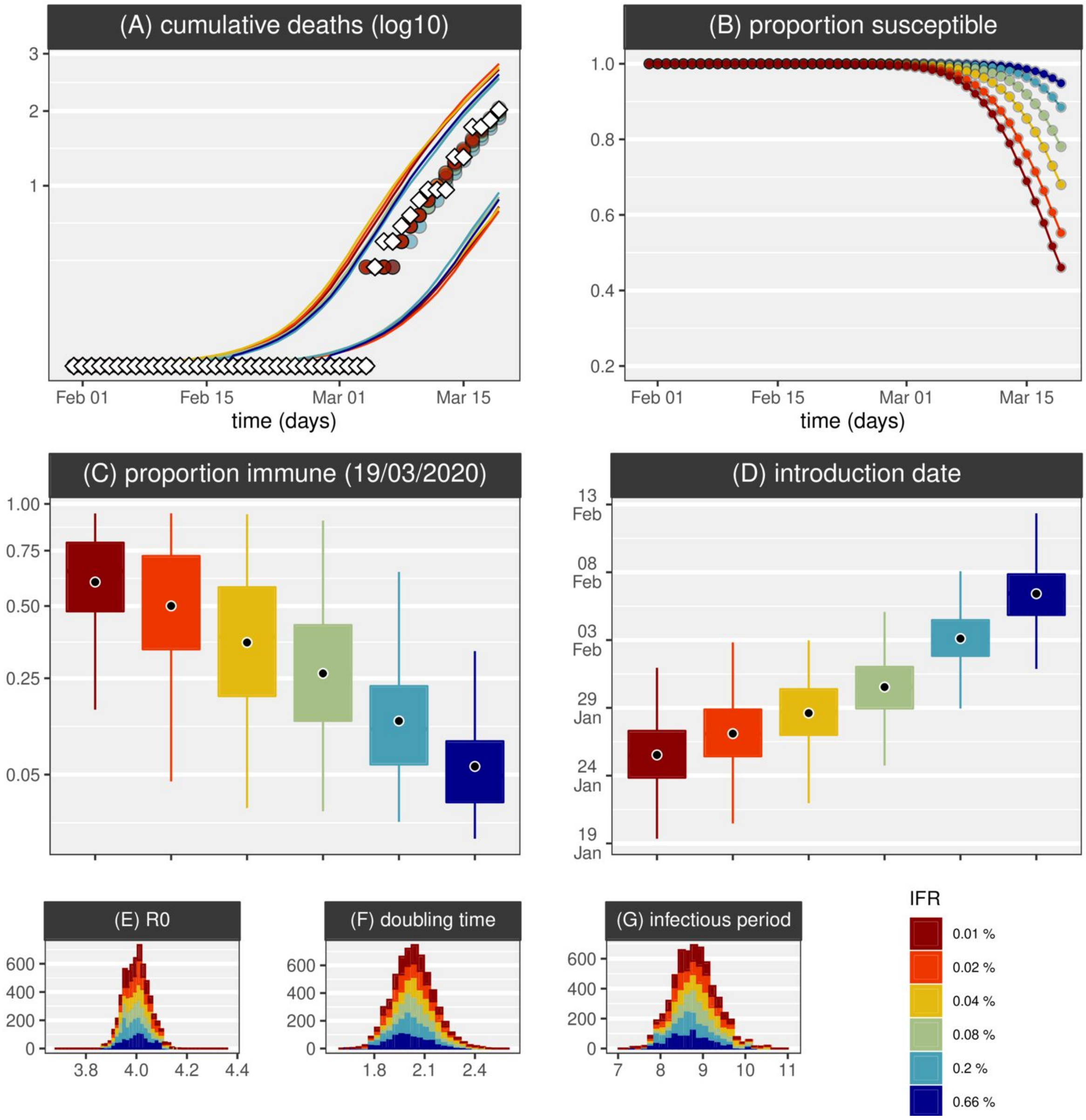


**Extra Figure 2 - SIR model, UK, single introduction,** $R_0 = 4$ **.** Figure legend the same as Figure 1 main text.

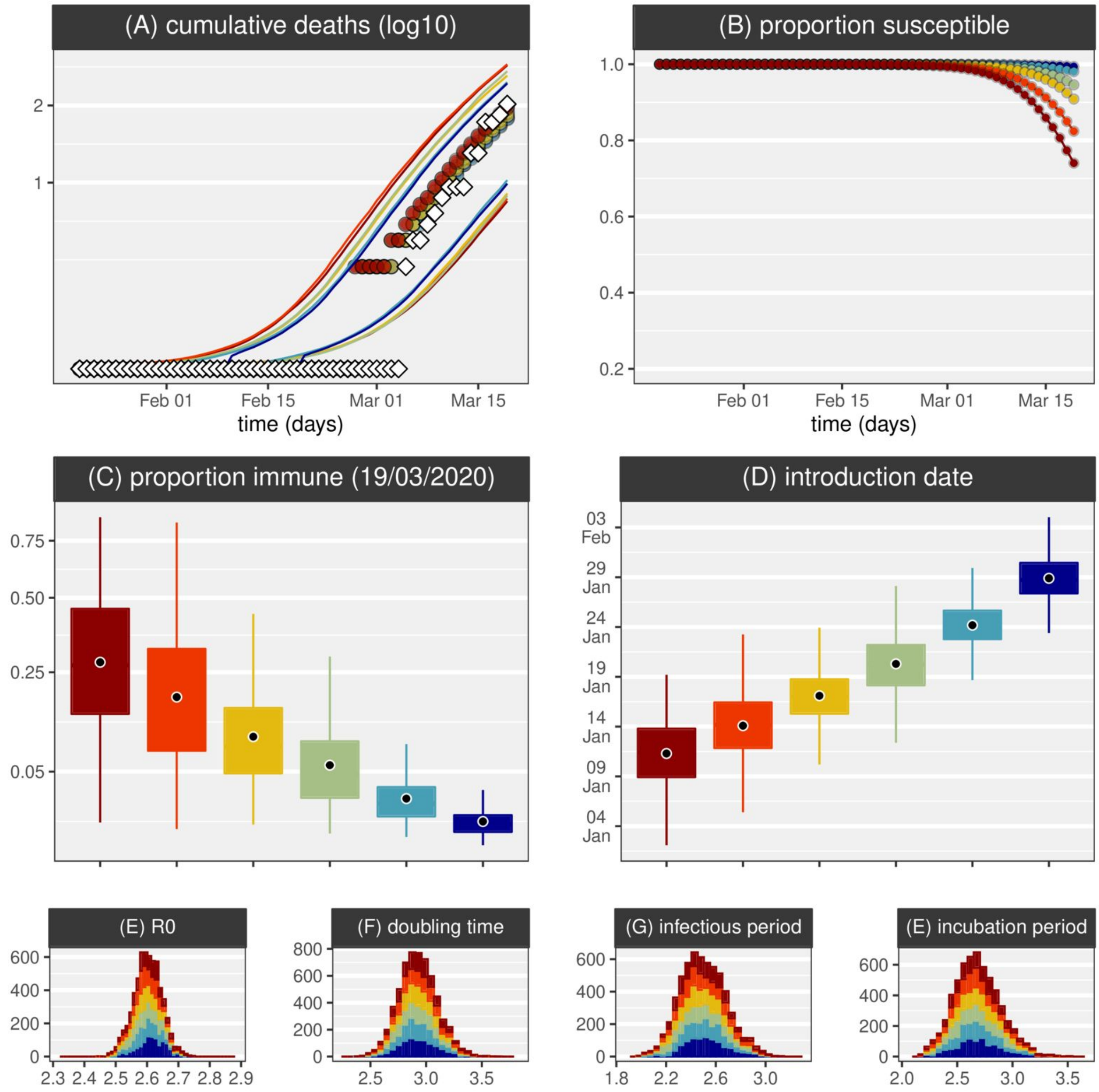


**Extra Figure 3 - SEIR model, UK, single introduction,** $R_0 = 2.5$ **.** Figure legend the same as Figure 1 main text.

We implemented SEIR by introducing an extra variable *e* to represent the exposed class and the following parameters:

| | | | |
|---|---|---|---|
| incubation period (days) | $1/\gamma$ | Gaussian distribution 1/G1(M=1/5,SD=0.05) | 41,45,46 |
| time (days) between symptom onset and death | $\psi$ | Gaussian distribution G1(M=12,SD=1.5) | 45 |

**equation 1a**: $de/dt \ = \beta y\ (1-z)\ -\ \gamma e$
**equation 2a**: $dy/dt \ = \gamma e\ -\sigma y$
**equation 3a**: $dz/dt \ = \beta y\ (1-z)$

**equation 4a**: $\Lambda_t \ = \eta\ N\ z_{t-\psi}$

**equation 5a**: $R_0 = \beta/(\sigma+\gamma)$

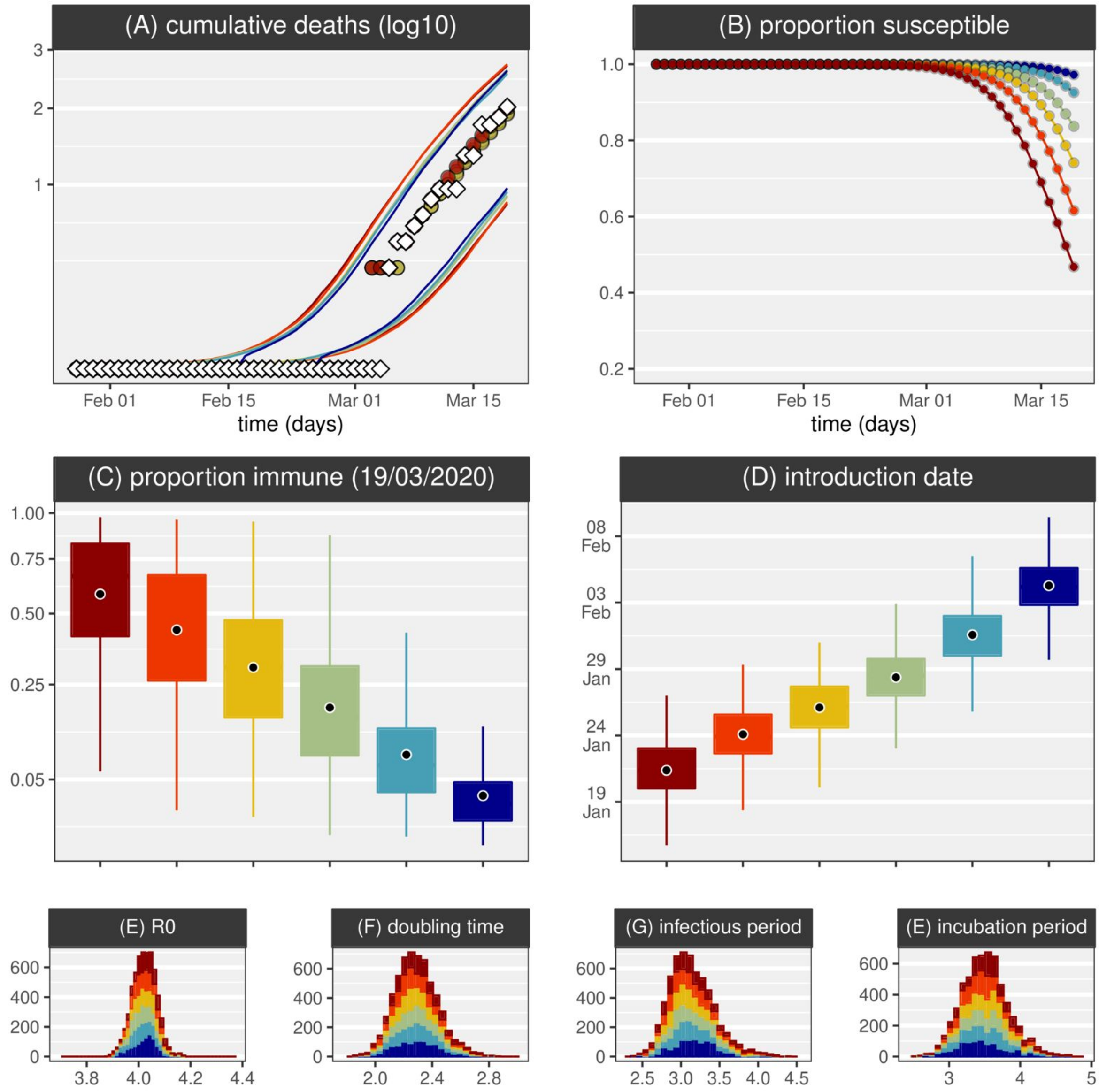


**Extra Figure 4 - SEIR model, UK, single introduction,** $R_0 = 4$. Figure legend the same as Figure 1 main text. Model details identical to Extra Figure 3.

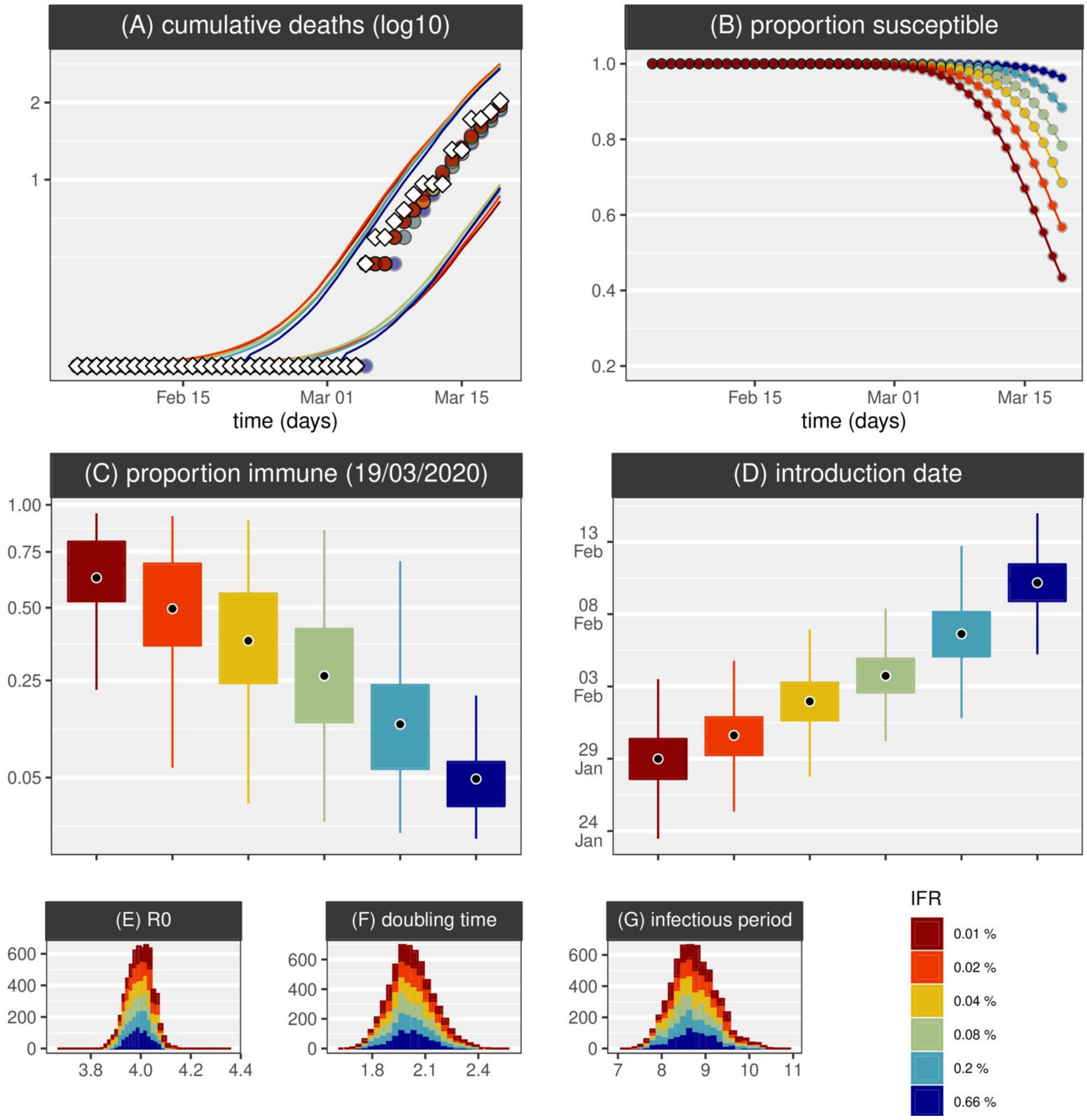


**Extra Figure 5 - SIR model, UK, with influx of infected,** $R_0 = 4$ **.** Figure legend the same as Figure 1 main text.

We model an external contribution to the force of infection growing at an exponentially rate (with a doubling time of 5 days) starting at estimated time of introduction $T_i$ (similar to [38]); stopping on 24/03/2020 for the UK and 11/03/2020 for IT (according to lockdown dates [31]). This external forcing was modelled by changing the term $\beta i\,(1-z)$ in **equations 1, 2** (SIR model) or **equations 1a, 3a** (SEIR model) to $\beta(i+m)\,(1-z)$, with $m = e^{t\tau}/N$. Model variables are summarized in **Table 1**, except:

| | | | |
|---|---|---|---|
| introduction external forcing growth rate | $\tau$ | 0.1386 | [38] |

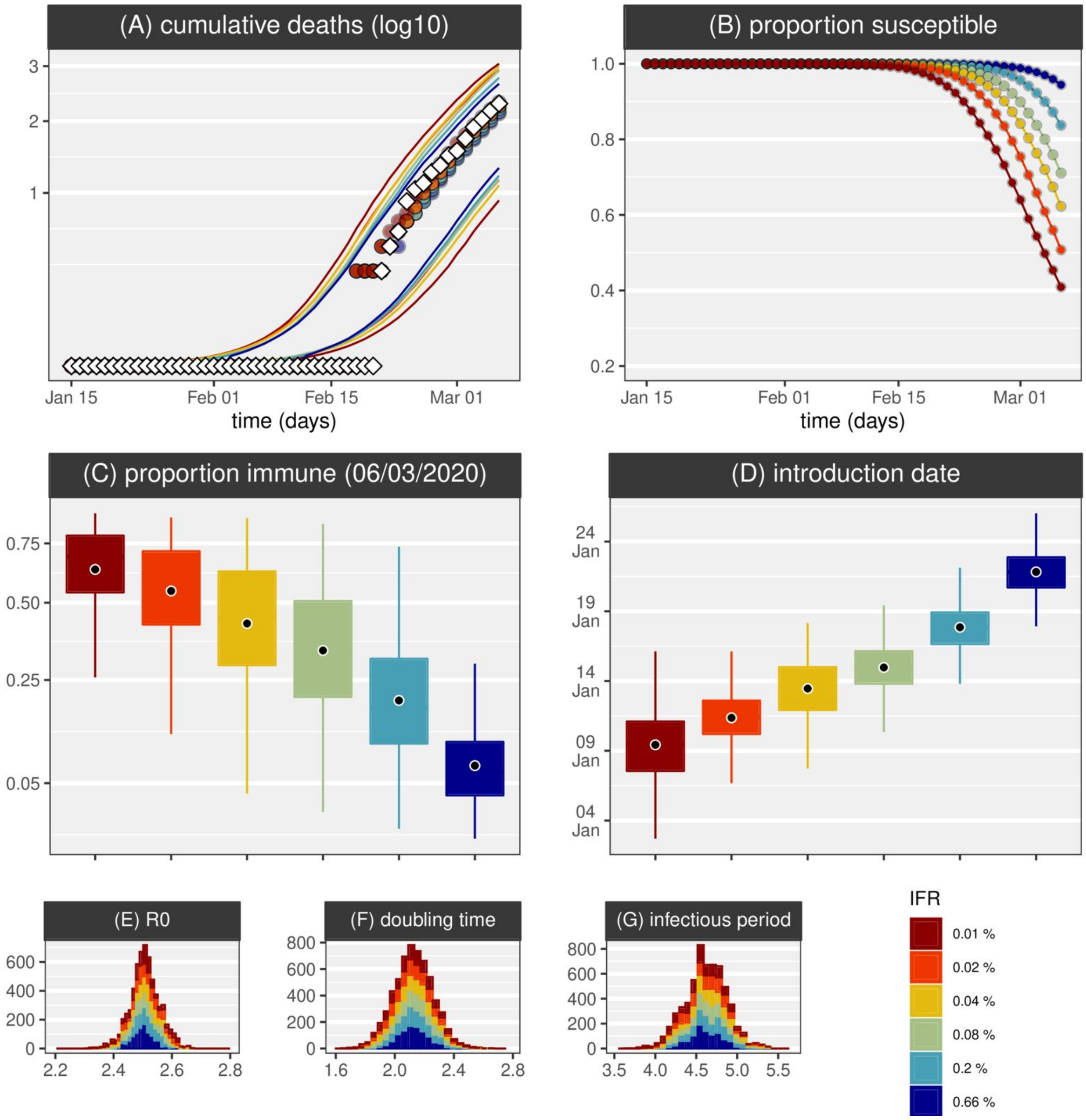


**Extra Figure 6 - SIR model, IT, single introduction,** $R_0 = 2.5$ **.** Figure legend the same as Figure 1 main text.

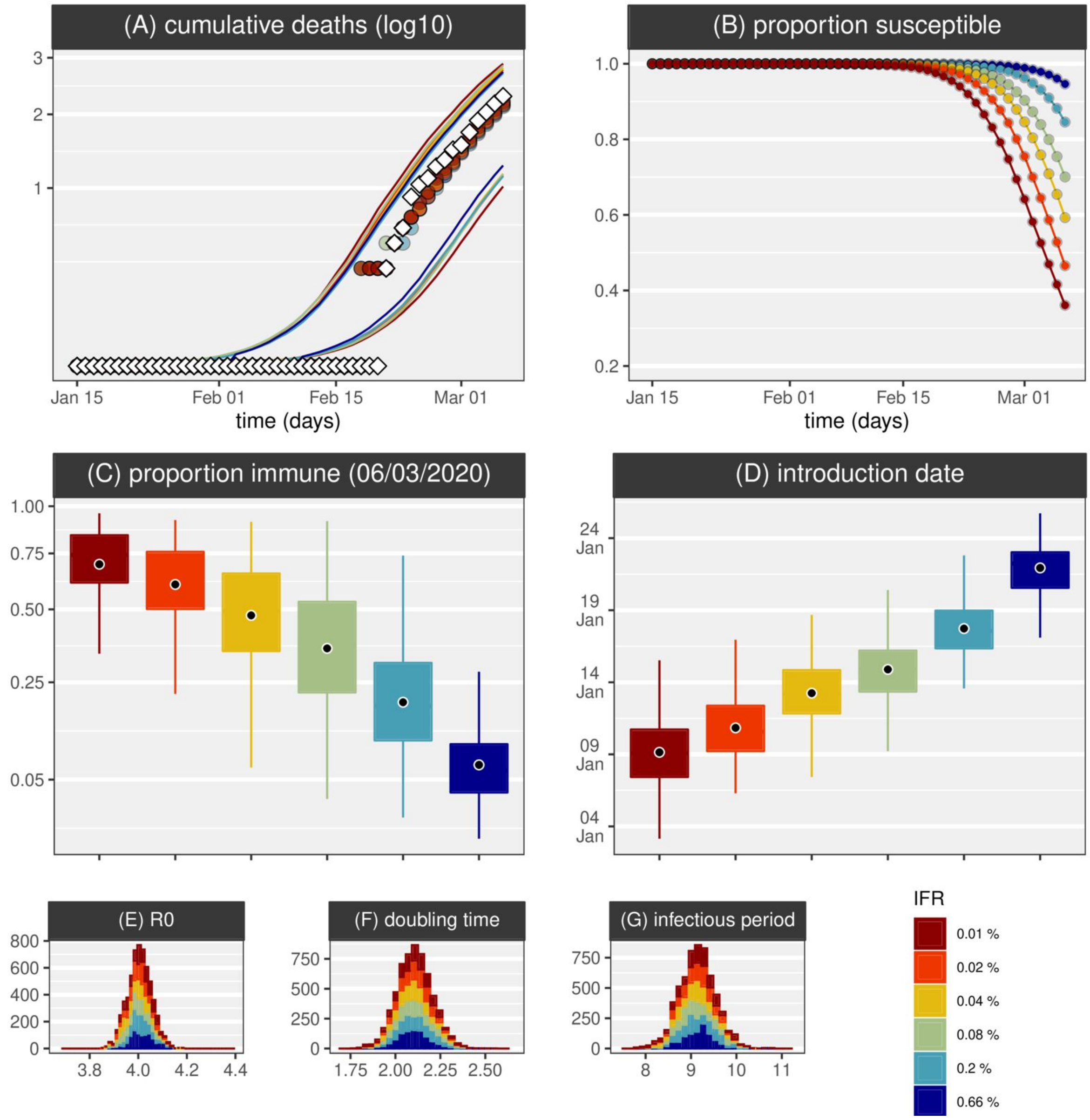


**Extra Figure 7 - SIR model, IT, single introduction,** $R_0 = 4$ **.** Figure legend the same as Figure 1 main text.

### References


1. Report of the WHO-China Joint Mission on Coronavirus Disease 2019 (COVID-19). 16-24 February 2020. Available: https://www.who.int/docs/default-source/coronaviruse/who-china-joint-mission-on-covid-19-final-report.pdf

2. Yang J, Zheng Y, Gou X, Pu K, Chen Z, Guo Q, et al. Prevalence of comorbidities in the novel Wuhan coronavirus (COVID-19) infection: a systematic review and meta-analysis. Int J Infect Dis. 2020. doi:10.1016/j.ijid.2020.03.017

3. Zhou F, Yu T, Du R, Fan G, Liu Y, Liu Z, et al. Clinical course and risk factors for mortality of adult inpatients with COVID-19 in Wuhan, China: a retrospective cohort study. The Lancet. 2020. doi:10.1016/s0140-6736(20)30566-3

4. Verity R, Okell LC, Dorigatti I, Winskill P, Whittaker C, Imai N, et al. Estimates of the severity of coronavirus disease 2019: a model-based analysis. The Lancet Infectious Diseases. 2020. doi:10.1016/s1473-3099(20)30243-7

5. Flaxman S, Mishra S, Gandy A, Unwin HJT, Coupland H, Mellan TA, et al. Estimating the number of infections and the impact of non- pharmaceutical interventions on COVID-19 in 11 European countries. Imperial College London; 2020 Mar.

6. Sutton D, Fuchs K, D'Alton M, Goffman D. Universal Screening for SARS-CoV-2 in Women Admitted for Delivery. N Engl J Med. 2020. doi:10.1056/NEJMc2009316

7. Silverman JD, Hupert N, Washburne AD. Using ILI surveillance to estimate state-specific case detection rates and forecast SARS-CoV-2 spread in the United States. doi:10.1101/2020.04.01.20050542

8. Boëlle P-Y, Souty C, Launay T, Guerrisi C, Turbelin C, Behillil S, et al. Excess cases of influenza-like illnesses synchronous with coronavirus disease (COVID-19) epidemic, France, March 2020. Eurosurveillance. 2020. doi:10.2807/1560-7917.es.2020.25.14.2000326

9. Jombart T, van Zandvoort K, Russell T, Jarvis C, Gimma A, Abbott S, et al. Inferring the number of COVID-19 cases from recently reported deaths. doi:10.1101/2020.03.10.20033761

10. Liu T, Hu J, Xiao J, He G, Kang M, Rong Z, et al. Time-varying transmission dynamics of Novel Coronavirus Pneumonia in China. doi:10.1101/2020.01.25.919787

11. Read JM, Bridgen JRE, Cummings DAT, Ho A, Jewell CP. Novel coronavirus 2019-nCoV: early estimation of epidemiological parameters and epidemic predictions. doi:10.1101/2020.01.23.20018549

12. Ferguson NM, Laydon D, Nedjati-Gilani G, Imai N, Ainslie K, Baguelin M, et al. Impact of non-pharmaceutical interventions (NPIs) to reduce COVID- 19 mortality and healthcare demand. Imperial College London; 2020 Mar.

13. Russell TW, Hellewell J, Jarvis CI, van Zandvoort K, Abbott S, Ratnayake R, et al. Estimating the infection and case fatality ratio for coronavirus disease (COVID-19) using age-adjusted data from the outbreak on the Diamond Princess cruise ship, February 2020. Eurosurveillance. 2020. doi:10.2807/1560-7917.es.2020.25.12.2000256

14. Wu J, Leung K, Bushman M, Kishore N, Niehus R, Salazar P, et al. Estimating clinical severity of COVID-19 from the transmission dynamics in Wuhan, China. doi:10.21203/rs.3.rs-17453/v1

15. Anderson RM, Anderson B, May RM. Infectious Diseases of Humans: Dynamics and Control. Oxford University Press; 1992.

16. Lourenço J, Obolski U, Swarthout TD, Gori A, Bar-Zeev N, Everett D, et al. Determinants of high residual post-PCV13 pneumococcal vaccine-type carriage in Blantyre, Malawi: a modelling study. BMC Med. 2019;17: 219.

17. McNaughton AL, Lourenço J, Hattingh L, Adland E, Daniels S, Van Zyl A, et al. HBV vaccination and PMTCT as elimination tools in the presence of HIV: insights from a clinical cohort and dynamic model. BMC Med. 2019;17: 43.

18. Lourenço J, de Lima MM, Faria NR, Walker A, Kraemer MUG, Villabona-Arenas CJ, et al. Epidemiological and ecological determinants of Zika virus transmission in an urban setting. eLife. 2017. doi:10.7554/elife.29820

19. Faria NR, da Costa AC, Lourenço J, Loureiro P, Lopes ME, Ribeiro R, et al. Genomic and epidemiological characterisation of a dengue virus outbreak among blood donors in Brazil. Sci Rep. 2017;7: 15216.

20. Kucharski AJ, Russell TW, Diamond C, Liu Y, Edmunds J, Funk S, et al. Early dynamics of transmission and control of COVID-19: a mathematical modelling study. Lancet Infect Dis. 2020. doi:10.1016/S1473-3099(20)30144-4

21. Park SW, Cornforth DM, Dushoff J, Weitz JS. The time scale of asymptomatic transmission affects estimates of epidemic potential in the COVID-19 outbreak. medRxiv. 2020. doi:10.1101/2020.03.09.20033514

22. Hellewell J, Abbott S, Gimma A, Bosse NI, Jarvis CI, Russell TW, et al. Feasibility of controlling COVID-19 outbreaks by isolation of cases and contacts. Lancet Glob Health. 2020;8: e488–e496.

23. Hilton J, Keeling MJ. Estimation of country-level basic reproductive ratios for novel Coronavirus (COVID-19) using synthetic contact matrices. doi:10.1101/2020.02.26.20028167

24. Linton NM, Kobayashi T, Yang Y, Hayashi K, Akhmetzhanov AR, Jung S-M, et al. Incubation Period and Other Epidemiological Characteristics of 2019 Novel Coronavirus Infections with Right Truncation: A Statistical Analysis of Publicly Available Case Data. J Clin Med Res. 2020;9. doi:10.3390/jcm9020538

25. Li Q, Guan X, Wu P, Wang X, Zhou L, Tong Y, et al. Early Transmission Dynamics in Wuhan, China, of Novel Coronavirus-Infected Pneumonia. N Engl J Med. 2020. doi:10.1056/NEJMoa2001316

26. Woelfel R, Corman VM, Guggemos W, Seilmaier M, Zange S, Mueller MA, et al. Clinical presentation and virological assessment of hospitalized cases of coronavirus disease 2019 in a travel-associated transmission cluster. doi:10.1101/2020.03.05.20030502

27. COVID-19 Italia - Monitoraggio situazione; Presidenza del Consiglio dei Ministri - Dipartimento della Protezione Civile. Available: https://github.com/pcm-dpc/COVID-19

28. Novel Coronavirus (COVID-19) Cases, provided by CSSE at Johns Hopkins University. Available: https://github.com/CSSEGISandData/COVID-19
