## Supplementary material for "The impact of host resistance on cumulative mortality and the threshold of herd immunity for SARS-CoV-2": Figures

### Supplementary Figures & Tables

| | **Proportionate mixing** ($\rho = \delta$) | | | | | **Assortative mixing** ($\delta = 1$) | | | | |
|---|---|---|---|---|---|---|---|---|---|---|
| | $R_0 = 1.25$ | $R_0 = 1.5$ | $R_0 = 2$ | $R_0 = 2.5$ | $R_0 = 3$ | $R_0 = 1.25$ | $R_0 = 1.5$ | $R_0 = 2$ | $R_0 = 2.5$ | $R_0 = 3$ |
| $\rho = 0$ | 0.2 | 0.33 | 0.5 | 0.6 | 0.66 | 0.2 | 0.33 | 0.5 | 0.6 | 0.66 |
| $\rho = 0.1$ | 0.1 | 0.23 | 0.4 | 0.5 | 0.56 | 0.18 | 0.3 | 0.45 | 0.54 | 0.6 |
| $\rho = 0.2$ | 0 | 0.13 | 0.3 | 0.4 | 0.46 | 0.16 | 0.26 | 0.4 | 0.48 | 0.53 |
| $\rho = 0.3$ | 0 | 0.03 | 0.2 | 0.3 | 0.36 | 0.14 | 0.23 | 0.35 | 0.42 | 0.46 |
| $\rho = 0.4$ | 0 | 0 | 0.1 | 0.2 | 0.26 | 0.12 | 0.2 | 0.3 | 0.36 | 0.4 |
| $\rho = 0.5$ | 0 | 0 | 0 | 0.1 | 0.16 | 0.1 | 0.16 | 0.25 | 0.3 | 0.33 |

**Table S1 - Herd-immunity threshold (HIT).** Model output for proportionate mixing ($\rho = \delta$, left, light grey) and assortative mixing ($\delta = 1$, right, white) for varying basic reproduction number $R_0$, proportion resistant $\rho$, and $R_{0_1} = 0$, $R_{0_2} = R_0$.

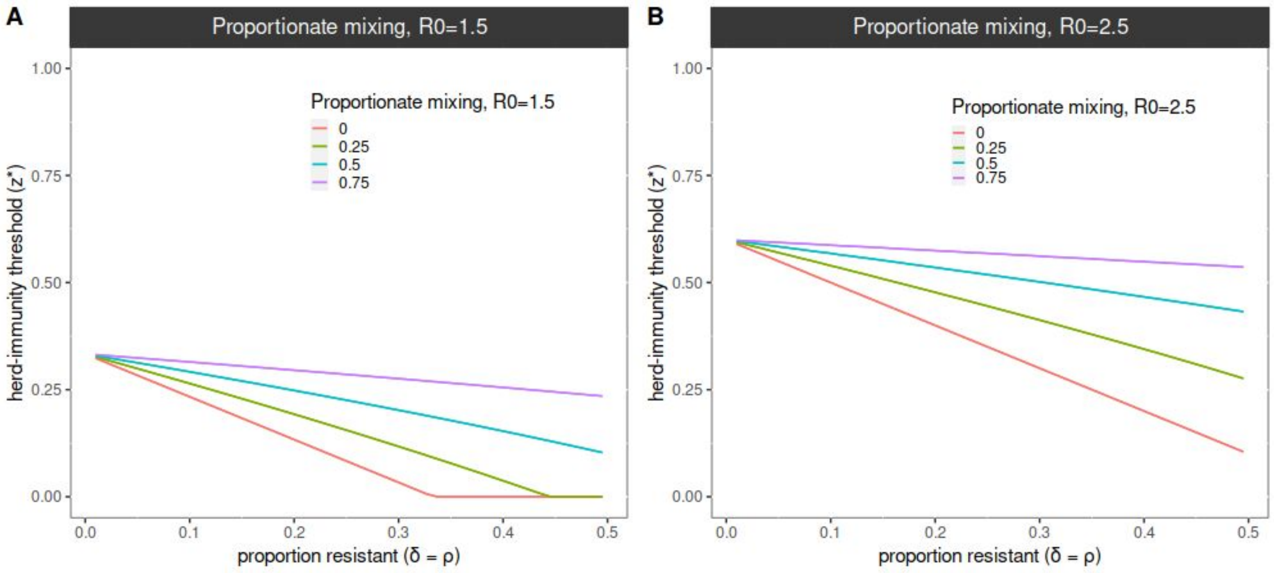


**Figure S1 - Herd-immunity threshold under proportionate mixing and incomplete resistance.** (left) Herd-immunity threshold (z*) under proportionate mixing ($\delta = \rho$) and incomplete resistance $F = R_{0_1}/R_{0_2}$ with $R_0 = 1.5$. (right) Same output as in the left panel but for with $R_0 = 2.5$. The scenario of $F = 0$ is fully explored in the main text. Simulations ran for 365 days with an infectious period ($1/\sigma$) of 5 days, $\gamma = 0$, $\delta = \rho$.

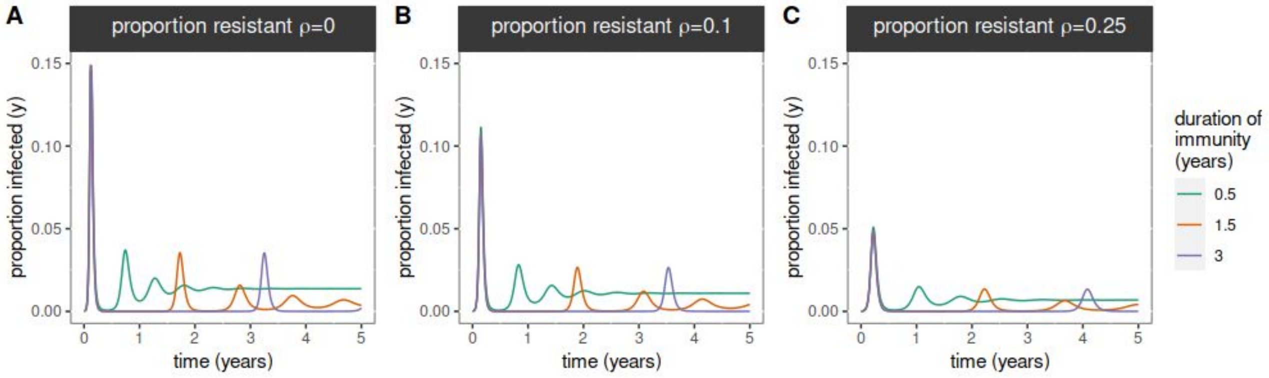


**Figure S2 - Effect of duration of immunity.** Time series of proportion infected (y) for varying proportion of resistant ( $\rho$ ) and duration of immunity ( $1/\gamma$ , color scale) under proportionate mixing ( $\delta = \rho$ ). Proportion of resistant ( $\rho$ ) for each panel is presented on the title. Infectious period $1/\sigma =$ 5 days.
